# Evaluation of the Nutritional Quality of Ultra-Processed Foods (Ready to Eat + Fast Food): Fatty Acid Composition

**DOI:** 10.1101/2021.04.16.21255610

**Authors:** Lisaura Maldonado-Pereira, Carlo Barnaba, Gustavo De los Campos, Ilce Gabriela Medina-Meza

**Affiliations:** Department of Chemical Engineering and Materials Science, Michigan State University, East Lansing, MI, USA.; Institute of Quantitative Health Science and Engineering, Michigan State University, East Lansing, MI, USA.; Department of Epidemiology and Biostatistics, Michigan State University, East Lansing, MI, USA.; Department of Biosystems and Agricultural Engineering, East Lansing, MI, USA.

**Keywords:** Ultra-processed foods, fatty acids, nutritional quality, salt, public health

## Abstract

Up to the 60% of the Western diet is characterized by consumption of high amounts of Ultra-processed foods (UPFs). From a nutritional standpoint, the high consumption of UPFs, including Fast-foods and Ready-to-Eat (RTE), emerges as a critical topic for public health linking nutritional quality and food safety. In the present work, we provide a systematic database of fatty acids composition of the most consumed UPFs in the US mid-western region. Saturate (SFA) and monounsaturated (MUFA) fatty acids were predominant in both FF and RTE, while health-beneficial polyunsaturated (PUFA) fatty acids were abundant only in seafood meals. Nutritional and non-nutritional attributes were correlated, finding positive correlation between caloric and sodium content. The significance of this study relies on providing new quantitative data for the most consumed UPFs, with the opportunity to define priority interventions for more advanced precision nutrition, especially for vulnerable populations.

## Introduction

Dietary lipids provide up to 42% of the calories ingested in the Western diet, whereas nutritional recommendations of fat for adults are 20–35% (Niot, Poirier, Tran, & Besnard, 2009). In the Western diet, ∼95% of dietary lipids are comprised of triacylglycerols, (mainly long-chain fatty acids (LCFA)), and the remaining includes phospholipids (4.5%) and cholesteryl esters (0.5%). Adherence to a “Westernized” dietary pattern is characterized by the consumption of high amounts of processed meats (Hu, 2002; Schulze & Hu, 2002). Nearly 60% of the calories consumed in the Western diet come from the intake of ultra-processed foods (UPFs) (Martínez Steele et al., 2016; Urban et al., 2016). Fast foods (FF) (including small, large, and non-chain restaurants) and Ready to Eat (RTE) meals constitute the people’s top choices. A healthy human with an average of 70 kg of body weight, stores around 141,000 kcal as fat as compared to 24,000 kcal as protein and 1,000 kcal as carbohydrate (Wang, Liu, Portincasa, & Wang, 2013). Furthermore, dietary fat is the most calorically dense macronutrient, supplying 9 kcal/g, about double of what is contributed by either protein or carbohydrate at 4 kcal/g. The increase of consumption of dietary fat from UPFs associated with a qualitative imbalance (excess of saturated fatty acids and cholesterol) has been associated with the increased risk in the development of several chronic diseases such as obesity, diabetes, cancer, and cardiovascular disease (Zinocker & Lindseth, 2018).

Food processing does not represent an issue for human nutrition, neither does it inherently cause negative health outcomes. However, the over-processing of food can lead to the accumulation of harmful substances formed because of thermal treatments, light exposure, storage, and aging, the denaturalization of proteins, vitamins, and bioactive components, compromising the overall nutritional quality.

UPFs are food items considered inexpensive, *highly processed*, rich in calories, but low in some essential micronutrients such as minerals and vitamins. UPFs are mostly consumed away from home and have been proposed as a major contributor to the energy intake rise (Urban et al., 2016). A big group of meals highly consumed among the UPFs is the ready-to-eat (RTE) meals. RTE meals are defined as any food that is either normally eaten in its raw state or has been processed, for which it is reasonably foreseeable that the food will be eaten without further processing that would significantly minimize biological hazards (FDA, 2020c). Usually, RTE meals are packed processed products for sale which require minimum preparation at home.

Most foods from the Western diet, including FFs and RTE meals, belong to UPFs category according to the NOVA (*not an acronym*) classification system (Monteiro, Cannon, Lawrence, Costa Louzada, & Pereira Machado, 2019). These terms differ in the number of processing techniques applied during the production of the food item. NOVA is recognized by the Food and Agriculture Organization (FAO) and Pan-American Health Organization (PAHO) as a valid tool for nutrition and public health research and “categorizes foods according to the extent and purpose of food processing, rather than in terms of nutrients.” (Juul et al., 2018; Martínez Steele et al., 2016; Moubarac et al., 2017). Since, consumers are continuously exposed to “new” UPFs with more than 96% of persuasive advertisement strategies (Santana et al., 2020), from a nutritional perspective, over-processing and chemical risk emerge as a critical topic for public health linking nutritional quality and food safety.

This study aimed to determine the fat composition of a selection of the most consumed UPFs in the Mid-West market. UPFs were analyzed based on fat sources, price, category of food, sugars, salt, and calorie content. This study will establish a database useful not only for nutritional and clinical interventions but also to improve food chemical safety and nutritional food quality.

## 2. Materials and Methods

### 2.1. Materials, Chemicals and Reagents

Methanol was from Sigma-Aldrich (St. Louis, MO). Chloroform was obtained from Omni Solv (Burlington, MA), hexane was purchased from VWR BDH Chemicals (Batavia, IL), 1-butanol, and potassium chloride (KCl) from J. T. Baker (Allentown, PA), and diethyl ether was purchased from Fisher Chemical (Pittsburgh, PA). Sodium sulfate anhydrous (Na_2_SO_4_) and sodium chloride (NaCl) were also purchased from VWR BDH Chemicals. Supelco 37 FAME standard mixture was purchased from Sigma Aldrich (St Louis, MO).

### 2.2. Sample collection

Composite foods and food products of different categories of UPFs were collected from retail stores, supermarkets, food chains, restaurants, and takeaway in the Lansing area (Michigan, USA) between February 2018 and October 2019. Different brands and different retailers for the same type of food were acquired to achieve a representative sample. A complete list of the food meals, their respective test code, and the group is provided in Supplemental Information (**Table S1)**. Meals were classified into two main groups: Fast Food (FF) and Ready to Eat (RTE), where 23 FF meals were purchased from the eight most popular franchises in the state of Michigan covering more than 75% of the national market (Dunford et al., 2017; Powell et al., 2019; Rwithley, 2019; Tran et al., 2019). Also, food items and meals were grouped in subcategories according to the fat source as follows: eggs and egg’s derivatives (E), dairy products (D), meat and poultry (MP), seafood (S), baby food (BF). Additional food items that did not fit in any of the previous categories, such as potato-products (potato crisps with and without added flavors, French fries from restaurants and takeaway, frozen potatoes pre-fried and fried, and homemade French fries), pasta, salad dressings, and popcorn (sweet or salty) were grouped as other products (O). Once the UPF arrived at the laboratory, an excel form was filled out with the following information: (1) UPF’s name; (2) price; (3) place of purchase, date, and time of collection; (4) type of food (RTE or FF); (5) nutritional declaration (energy, fat, saturated fatty acids, carbohydrates, sugars, fiber, protein, and salt); (6) portion size; (7) list of ingredients; (8) expiring date; and (9) other relevant information.

### 2.3. Sample preparation

FF meals were purchased from several franchises and brought to the laboratory for immediate analysis. RTE meals were purchased from different local supermarket stores, and immediately brought to the laboratory. Storage conditions were followed according to the label instructions (fresh foods were kept in a fridge at 4°C and frozen meals were kept at -20°C or the temperature indicated in the label). All UPFs were analyzed before the expiration date. When further cooking procedures were required, items were prepared following manufacturer’s instructions. Kitchen equipment available at the Michigan State University’s Food Science Laboratory was supplied and used for this study. All the samples were homogenized using an Ultra-Turrax® (Tekmar TP 18/10S1 Cincinnati, OH) for 3 min at 5000 rpm, split, and stored accordingly, depending on the food matrix.

### 2.4. Lipid Extraction

Lipid fraction was extracted according to the Folch cold extraction method (Folch, Lees, & Sloane Stanley, 1957) with some modifications depending on the food matrix. Thirty grams of sample were minced and placed in a 500 mL glass bottle with a screwcap where 200 mL of chloroform:methanol solution (1:1, v/v) was added. The sample was mixed for 15 min at 300 rpm. Homogenization was performed using an Ultra-Turrax for 3 minutes. The bottle was kept in an oven at 60°C for 20 min before adding 100 mL chloroform. After 2 min of vortex mixing the sample, the content of the bottle was filtered. The filtrate was mixed thoroughly with 100 mL of 1 M KCl solution, and left overnight at 4 °C. Then, the lower phase containing lipids was collected and dried at 60°C with a multi-vacuum solvent evaporator (Organomation S-EVAP-RB, Berlin MA) at 25 inches of Hg. Total fat content was determined gravimetrically.

### 2.5. Fatty acid methyl esters (FAME)

Fatty acid methyl esters (FAME) were prepared according to the transesterification described in Chen’s procedure (Chen, Aluwi, Saunders, Ganjyal, & Medina-Meza, 2019). One μL of the methylated sample was injected into a gas chromatograph (GC 2010 Shimadzu, Kyoto Japan) equipped with a DB-WAX capillary column (30 m × 0.32 mm i.d. × 0.25 μm). The oven temperature gradient was set as follows: 120 °C to 200°C with a rate of 3°C/min, then from 200 °C to 240°C with a rate of 2 °C/min and held for 2 min. Injector and detector were both set at 250°C. H_2_ was used as carrier gas at 1mL/min and split ratio 5.0. Data acquisition was done by Lab solutions software (Shimadzu, Kyoto, Japan) and peak areas were identified by comparing their retention times to the pure standards. Standard curves of FAME 37 mixture were built with different concentrations from 50 to 500 ug/mL. Fatty acid contents were reported as the weight percentage of total fatty acid detected (% w/w) per g of fat.

Results on fatty acids contents were obtained using the sums: Σ saturated fatty acids (Σ SFA = C8:0 + C10:0 + C11:0 + C12:0 + C13:0 + C14:0 + C15:0 + C16:0 + C17:0 + C18:0 + C20:0 + C21:0 + C22:0 + C23:0 + C24:0); Σ monounsaturated fatty acids (Σ MUFA = C14:1, cis-9 + C15:1, cis-10 + C16:1, cis-9 + C17:1, cis-10 + C18:1, cis-9 + C20:1, cis-11 + C22:1, cis-13 + C24:1, cis-15); Σ polyunsaturated fatty acids (Σ PUFA = C18:2, cis-9,12 + C18:3, cis-6,9,12 + C18:3, cis-9,12,15 + C20:2, cis-11,14 + C20:3, cis-8,11,14 + C20:3, cis-11,14,17 + C20:4, cis-5,8,11,14 + C22:2, cis-13,16 + C20:5, cis-5,8,11,14,17 + C22:6, cis-4,7,10,13,16,19). and Σ trans fatty acids (Σ TFA= C18:1, trans-9 + C18:2, trans-9,12).

### 2.5 Statistical analysis

Descriptive statistics were calculated overall and by category (RTE/FF) and food origin (meat, dairy, baby foods, seafood, egg and eggs products, others). Both mean, and confidence interval (95%) were computed. Since the data did not follow a normal distribution, when comparing RTE vs FF items, a Matt-Whitney *U-*test was performed, at *p* < 0.05 significance level. Statistical differences between food categories were evaluated by using the non-parametric Kruskal-Wallis ANOVA by Ranks test, at *p* < 0.05 significance level. Spearman’s correlation across variables was also tested. All the statistical analyses were computed using SPSS v.27 (IBM).

## Results and discussion

### 3.1 Total Fat, Sugar and Sodium in UPFs

The overall total fat ranges from 0.60 to 87.62 g/100 g of product with the dairy category (n=11) being the group with the highest fat content followed by the eggs and egg’s derivatives category (n=2) with up to 77 g/100 g of product (**Table 1**).

**Table 1.** Food ID, Total fat, Sugar, Sodium and Calories per serving found in the different UPFs.

| Category | Food ID | Total Fat<br>(g/100 g sample) | Total Fat<br>Nutritional<br>label **<br>(g/100 g<br>sample) | Serving Size | Total Fat (g)<br>per serving<br>size | Sugar (g)<br>per serving<br>size | Sodium<br>(mg)<br>per<br>serving | Calories<br>per<br>serving |
| --- | --- | --- | --- | --- | --- | --- | --- | --- |
| Ready to Eat |  |  |  |  |  |  |  |  |
| Dairy | D1 - RTE | 17.67 ± 2.45 | 18.40 | 19.00 g | 3.50 | 1.00 | 280 | 60 |
|  | D2 - RTE | 26.92 ± 8.47 | 33.00 | 21.00 g | 6.93 | 0 | 180 | 110 |
|  | D3 - RTE | 49.71 ± 2.56 | 50.00 | 1 tbsp (14.15 g) | 7.08 | NR | 100 | 60 |
|  | D4 - RTE | 87.62 ± 1.91 | 78.57 | 14.00 g | 11.00 | 0 | 90 | 100 |
|  | D5 - RTE | 10.27 ± 0.38 | 10.00 | 30.00 mL (28.3 g) | 2.83 | 1 | 25 | 40 |
|  | D6 - RTE | 24.16 ± 6.98 | 28.00 | 28.00 g | 7.84 | 1 | 240 | 60 |
|  | D7 - RTE | 42.82 ± 1.68 | 35.00 | 28.00 g | 9.80 | 2 | 95 | 90 |
|  | D8 - RTE | 12.78 ± 3.04 | 23.40 | 0.50 cup (65.00 g) | 15.21 | 14 | 40 | 130 |
|  | D9 - RTE | 1.60 ± 0.31 | 3.03 | 1 container (99.00g) | 3.00 | 20 | 90 | 150 |
|  | D10 - RTE | 2.14 ± 0.54 | 0.96 | 414.03 g | 0.60 | 37 | 230 | 250 |
|  | D11 - RTE | 12.56 ± 0.38 | 0.69 | 1 scoop (9.00 g) | 0.062 | NR | 0.32 | 44 |
| Meat &<br>Poultry | MP1 - RTE | 23.45 ± 5.00 | 20.00 | 38.00 g | 7.60 | < 1 | 260 | 90 |
|  | MP2 - RTE | 17.84 ± 2.65 | 26.00 | 32.00 g | 8.30 | 0 | 260 | 50,000 |
|  | MP3 - RTE | 0.60 ± 0.071 | 0.65 | 240.00 mL (226.40 g) | 1.50 | 3 | 870 | 170 |
|  | MP4 - RTE | 3.64 ± 0.018 | 3.30 | 1 can (236.59 g) | 7.80 | 5 | 870 | 250 |
|  | MP5 - RTE | 3.53 ± 0.55 | 5.65 | 11.00 oz (227.00 g) | 12.80 | 5 | 500 | 270 |
|  | MP6 - RTE | 1.37 ± 0.31 | 1.76 | 0.5 cup (113.40 g) | 2.00 | 0 | 640 | 60 |
|  | MP7 - RTE | 3.61 ± 0.013 | 0.61 | 0.5 cup (240.00 g) | 1.50 | 1 | 600 | 70 |
|  | MP8 - RTE | 2.91 ± 0.53 | 2.87 | 1 cup (249.00 g) | 7.15 | 6 | 800 | 200 |
|  | MP9 - RTE | 4.61 ± 0.097 | 4.28 | 1 cup (257.00 g) | 11.00 | 8 | 800 | 260 |
| Seafood | S1 - RTE | 3.62 ± 0.25 | 3.38 | 18.80 oz (532.97 g) | 18.01 | 1 | 890 | 180 |
| Egg's<br>Derivatives | E1 - RTE | 77.68 ± 1.95 | 75.00 | 1 Tbsp (13 g) | 9.75 | NR | 90 | 90 |
|  | E2 - RTE | 14.64 ± 1.02 | 13.91 | ½ cup (115.00 g) | 16.00 | 10 | 330 | 380 |
| Baby Food | BF1 - RTE | 1.80 ± 0.15 | 1.41 | 71.00 g | 1.00 | 0 | 35 | 50 |
|  | BF2 - RTE | 7.28 ± 0.19 | 4.40 | 4.00 oz (113.40 g) | 4.99 | NS | 40 | 90 |
|  | BF3 - RTE | 2.99 ± 0.06 | 4.40 | 4.00 oz (113.40 g) | 4.99 | 3 | 30 | 70 |
|  | BF4 - RTE | 3.25 ± 0.23 | 4.40 | 4.00 oz (113.40 g) | 4.99 | 3 | 40 | 70 |
|  | BF5 - RTE | 3.23 ± 0.14 | 4.40 | 4.00 oz (113.40 g) | 4.99 | 3 | 45 | 80 |
|  | BF6 - RTE | 2.12 ± 0.60 | 1.56 | 128.00 g | 2.00 | 5 | 260 | 120 |
|  | BF7 - RTE | 3.09 ± 0.16 | 4.40 | 4.00 oz (113.40 g) | 4.99 | 5 | 20 | 80 |
|  | BF8 - RTE | 6.00 ± 0.14 | 4.90 | 71.00 g | 3.48 | 0 | 20 | 50 |
|  | BF9 - RTE | 2.79 ± 0.44 | 4.40 | 4.00 oz (113.40 g) | 4.99 | 11 | 50 | 100 |
|  | BF10 - RTE | 4.63 ± 0.48 | 4.40 | 4.00 oz (113.40 g) | 4.99 | 4 | 40 | 80 |
|  | BF11 - RTE | 3.89 ± 0.23 | 4.40 | 4.00 oz (113.40 g) | 4.99 | 4 | 80 | 120 |
|  | BF12 - RTE | 2.90 ± 0.38 | 4.40 | 4.00 oz (113.40 g) | 4.99 | 6 | 75 | 90 |
|  | BF13 - RTE | 1.39 ± 0.25 | 2.35 | 85.00 g | 2.00 | 1 | 170 | 80 |
| Other | O1 - RTE | 19.97 ± 1.85 | 26.60 | 2 tbsp (30.00 g) | 7.98 | 0 | 290 | 140 |
|  | O2 - RTE | 45.11 ± 6.32 | 50.00 | 2 tbsp (30.00g) | 15.00 | 1 | 230 | 140 |
|  | O3 - RTE | 8.70 ± 5.00 | 5.00 | 1 package (58.00 g) | 2.90 | 6 | 570 | 250 |
|  | O4 - RTE | 3.65 ± 1.51 | 5.00 | 2.5 oz (70.90 g) | 3.55 | 4 | 500 | 220 |
| **Fast Food** |  |  |  |  |  |  |  |  |
| Meat &<br>Poultry | MP10 - FF | 33.13 ± 5.12 | 10.00 | 95.00 g | 9.50 | NR | 510 | 250 |
|  | M11 - FF | 19.52 ± 1.46 | 20.00 | 4 pieces (64.00 g) | 12.80 | NR | 330 | 170 |
|  | MP12 - FF | 23.59 ± 3.40 | 14.00 | 119.00 g | 16.66 | NR | 720 | 300 |
|  | MP13 - FF | 9.83 ± 0.45 | 11.40 | 1 taco (102.00 g) | 11.63 | 1 | 306 | 269 |
|  | MP14 - FF | 11.06 ± 2.09 | 7.60 | 1 quesadilla (170.00 g) | 12.92 | 0 | 310 | 180 |
|  | MP15 - FF | 9.95 ± 0.67 | 9.90 | 1 burrito (140.00 g) | 13.86 | 1 | 1650 | 640 |
|  | MP16 - FF | 12.48 ± 0.30 | 12.21 | 1 piece (75.00 g) | 9.16 | 0 | 430 | 130 |
|  | MP17 - FF | 19.39 ± 2.55 | 18.60 | 1 piece (60.00 g) | 11.16 | 0 | 380 | 130 |
|  | MP18 - FF | 6.26 ± 1.21 | 4.58 | 5.40 oz (153.09 g) | 7.01 | 7.00 | 520 | 150 |
|  | MP19 - FF | 8.19 ± 0.78 | 10.80 | 5.70 oz (161.69 g) | 17.46 | 19.00 | 820 | 490 |
|  | MP20 - FF | 14.23 ± 2.82 | 10.50 | 1 sandwich (187.00 g) | 19.64 | 9.00 | 680 | 320 |
|  | MP21 - FF | 7.12 ± 1.18 | 6.51 | 1 sandwich (71.28 g) | 4.64 | 3 | 2370 | 810 |
|  | MP22 - FF | 10.74 ± 4.18 | 8.79 | 1 sandwich (94.61 g) | 8.32 | 2 | 1940 | 850 |
|  | MP23 – FF*** | 12.59 ± 1.35 | 11.50 | 1 slice (123.00 g) | 14.15 | 3 | 950 | 370 |
|  | MP24 - FF | 10.96 ± 2.70 | 11.50 | 1 slice (79.00 g) | 9.09 | 2.00 | 740 | 380 |
| Seafood | S2 - FF | 22.82 ± 1.07 | 14.08 | 1 sandwich (131.00 g) | 18.44 | NR | 580 | 380 |
|  | S3 - FF | 21.70 ± 0.15 | 13.10 | 5.00 oz (141.75 g) | 18.57 | 14.00 | 440 | 360 |
| Other | O5 - FF | 13.92 ± 1.58 | 20.00 | 1 medium serving<br>(117.00 g) | 23.40 | NR | 260 | 320 |
|  | O6 - FF | 16.27 ± 2.86 | 16.00 | 1 biscuit (76.00 g) | 12.16 | NR | 810 | 260 |
|  | O7 - FF | 6.29 ± 1.34 | 6.00 | 3 hotcakes (149.00 g) | 8.94 | NR | 550 | 580 |
|  | O8 - FF | 18.16 ± 0.75 | 17.30 | 1 biscuit (49.00 g) | 8.48 | 1 | 520 | 180 |
|  | O9 - FF | 15.96 ± 0.21 | 15.00 | 1 order (34.99 g) | 5.25 | 0 | 1100 | 320 |
|  | O10 - FF | 1.46 ± 0.52 | 3.11 | 1 order (16.85 g) | 0.52 | 0 | 520 | 130 |

\*\*Information found in the USDA Food Database. www.fdc.nal.usda.gov/fdc-app.html#/
\*\*\*Information found in Menuwithprice.com. Not available in neither USDA Food Database nor Marco's Pizza website.
NR = not reported
NS = not a significant source of the nutrient

**Figure 1A-D** shows a boxplot distribution of fat and FAME groups within each category. UPFs meals and food products were grouped based on their fat source as described above. Some food products like edible fats (olive, avocado, canola, etc.) were not included since they are considered foods themselves with no additional components. Due to the large number of UPFs in this study, total fat will be primarily discussed for each food category, detailing it for individual meals only when a distinct result is statistically significant. FF and RTE meals are known to be served in large portions, containing high levels of saturated fat and added sugars (Harris et al., 2013; Rosenheck, 2008). Most of these meals are prepared with several ingredients such as oils, eggs, high fat meats - all of them containing high-fat content themselves - resulting in a meal with both high-fat and high caloric content. It is well known that UPFs are rich in fat, sugar, and salt, additionally consumption of these macronutrients is high among the US population (USDA, 2020b). Four UPFs from the category of FF contain half or more mg of salt than the recommended daily value (FDA, 2020b), being meat the major component of the meal preparation. Sugar-sweetened beverages, snacks and desserts are the major sources of added sugars in UPFs. In this study, the only beverage included exceeding the recommended daily value (Health, 2020). Therefore, the reduction of fat, salt, and sugar is a challenge that should be addressed without affecting the sensorial properties of the meals. Also, their reduction should be promoted in any food/meal manufacture restaurants, food chains, and takeaway stores; thus, this study serves as a database of the current content of fat and its relationship with salt and sugar in most consumed UPFs.

**Figure 1.**
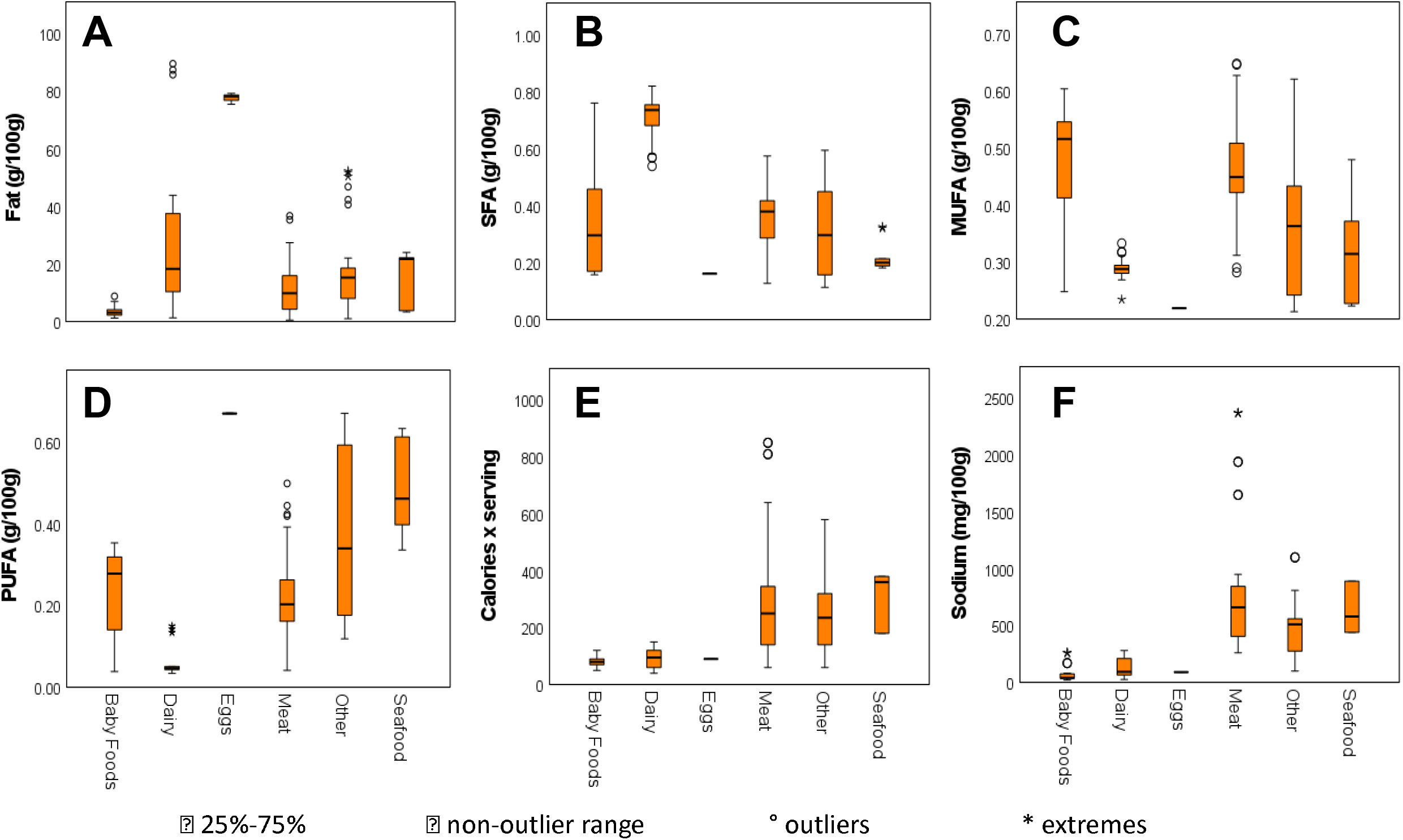
Total fat, SFA, MUFA, PUFA, Calories per serving and Salt (as mg/100 g of Sodium) of each food category

### 3.2 Fatty acid profile by food category

Thirty-five fatty acids were identified and quantified. Short-chain fatty acids (SC-FA) were not reported since C4 and C6 co-eluded with the solvent. Also, dihomo-γ-linolenic acid (C20:3n-3, DGLA) and eicosatrienoic acid (C20:3n-6) were not chromatographically resolved for some samples, therefore their area values were merged and reported as C20:3n-3 + C20:3n-6. **Table 2** and **Table 3** show the percentage of saturated fatty acids (SFA), monounsaturated fatty acids (MUFA), and polyunsaturated fatty acids (PUFA), respectively. For a better representation of the overall distribution of FAME s (SFA, MUFA, PUFA) according to the fat source a boxplot per food category is shown in **Figure 1**. No statistically significant differences were found between FF and RTE groups, however, differences among food categories were observed such as the meat and other categories.

**Table 2.** Saturated fatty acid composition in Ready to Eat and Fast Food meals.

| SFA | Total | Ready-To-Eat | Fast-Food | Mann-Whitney U-test |
| --- | --- | --- | --- | --- |
| **C8:0** | 1.18 (0.08-1.59) | 1.43 (0.08-2.03) | 0.64 (0.49-0.80) |  |
| **C10:0** | 1.74 (1.49-2.00) | 2.01 (0.17-2.36) | 1.22 (1.03-1.41) | * |
| **C11:0** | 0.06 (0.05-0.08) | 0.07 (0.06-0.09) | 0.04 (0.02-0.05) | *** |
| **C12:0** | 2.05 (1.63-2.46) | 2.04 (1.57-2.51) | 2.06 (1.22-2.89) |  |
| **C14:0** | 3.54 (2.99-4.09) | 4.22 (3.43-5.02) | 2.36 (1.81-2.91) | ** |
| **C15:0** | 0.42 (0.36-0.49) | 0.51 (0.42-0.06) | 0.28 (0.21-0.34) | * |
| **C16:0** | 21.74 (20.52-22.96) | 23.96 (22.34-25.57) | 18.09 (16.57-19.60) | **** |
| **C17:0** | 0.35 (0.31-0.39) | 0.39 (0.34-0.44) | 0.29 (0.22-0.35) | ** |
| **C18:0** | 7.61 (7.10-8.12) | 8.41 (7.69-9.13) | 6.29 (5.76-6.82) | *** |
| **C20:0** | 0.29 (0.26-0.32) | 0.24 (0.21-0.28) | 0.35 (0.31-0.40) | *** |
| **C21:0** | 0.30 (0.24-0.35) | 0.31 (0.22-0.40) | 0.27 (0.23-0.32) |  |
| **C22:0** | 0.12 (0.08 -0.15) | 0.12 (0.07-0.17) | 0.11<br>(0.06-0.17) |  |
| **C23:0** | 0.05 (0.03-0.08) | 0.03 (0.02-0.04) | 0.07 (0.03-0.10) | *** |
| **C24:0** | 0.22 (0.13-0.32) | 0.21 (0.06-0.35) | 0.24 (0.10-0.37) | *** |
| **Σ SFA** | 35.57 (33.01-38.13) | 40.88 (37.48-44.29) | 29.88 (27.17-32.60) | **** |
| **Σ MC-SFA** | 3.66 (2.98-4.34) | 3.94 (3.05-4.82) | 3.08 (2.06-4.10) |  |
| **Σ LC-SFA** | 33.66 (31.59-35.72) | 37.31 (34.48-40.15) | 27.61 (25.36-29.86) | **** |

ΣSFA: Sum of saturated fatty acid, ΣMC-FA: sum of medium chain saturated fatty acid, ΣLC-FA long chain saturated fatty acid.

**Table 3.** Monounsaturated and Polyunsaturated fatty acid composition in Ready to Eat and Fast Food meals

| MUFA | Total | Ready-To-Eat | Fast-Food | Mann-Whitney U-test |
| --- | --- | --- | --- | --- |
| **C14:1** | 0.55 (0.48-s0.62) | 0.59 (0.52-0.67) | 0.46 (0.32-0.60) | *** |
| **C15:1** | 0.18 (0.12-0.25) | 0.18 (0.12-0.25) | ND | - |
| **C16:1** | 1.76 (1.56-1.96) | 1.83 (1.57-2.09) | 1.63 (1.31-1.95) |  |
| **C17:1** | 0.26 (0.22-0.31) | 0.26 (0.20-0.31) | 0.28 (0.18-0.38) |  |
| ***t*C18:1** | 1.23 (0.77-1.70) | 2.14 (0.53-3.75) | 0.93 (0.59-1.27) |  |
| **C18:1 *n*-9** | 36.09 (34.65-37.53) | 33.64 (31.88-35.39) | 40.15 (37.92-42.38) | **** |
| *C20:1* | 0.40 (0.36-0.44) | 0.33 (0.28-0.39) | 0.49 (0.42-0.55) | **** |
| *C22:1* | 0.28 (0.20-0.36) | 0.19 (0.15-0.24) | 0.34 (0.21-0.47) | *** |
| *C24:1* | 0.19 (0.09-0.28) | 0.14 (0.05-0.23) | 0.27 (0.02-0.57) | *** |
| ***ΣMUFA*** | 37.82 (36.01-39.63) | 36.57 (34.64-38.50) | 43.17 (40.73-45.61) | **** |

| **PUFA** | **Total** | **Ready-To-Eat** | **Fast-Food** | **Mann-Whitney U-test** |
| --- | --- | --- | --- | --- |
| *tC18:2* | 0.35 (0.30-0.40) | 0.39 (0.31-0.46) | 0.31 (0.23-0.39) |  |
| ***C18:2 n-6*** | 20.88 (18.81-22.95) | 19.59 (16.88-22.29) | 23.02 (19.82-26.22) | * |
| *C18:3n-6* | 0.10- (0.08-0.12) | 0.11 (0.08-0.14) | 0.09 (0.07-0.10) |  |
| *C18:3n-3* | 2.70 (2.37-3.03) | 2.7 (2.21-3.14) | 2.75 (2.31-3.19) |  |
| *C20:3n3 + C20:3n6* | 0.12 (0.10-0.13) | 0.12 (0.10-0.15) | 0.11 (0.10-0.13) |  |
| *C20:4n-6* | 0.11 (0.05-0.17) | ND | 0.11 (0.05-0.17) |  |
| *C20:5n-3* | 0.27 (0.16-0.39) | 0.21 (0.12-0.30) | 0.32 (0.12-0.51) | *** |
| *C22:2 n-6* | 0.06 (0.00-0.13) | 0.02 (0.01-0.04) | 0.13 (-0.13-0.40) | **** |
| *C22:6 n-3* | 0.05 (0.04-0.07) | 0.05 (0.03-0.07) | 0.06 (0.04-0.07) | *** |
| ***ΣPUFA n-3*** | 0.17 (0.12-0.21) | 0.16 (0.10-0.21) | 0.18 (0.09-0.28) |  |
| ***ΣPUFA n-6*** | 23.88 (21.54-26.21) | 22.55 (19.46-25.63) | 26.07 (22.51-29.63) | * |
| ***ΣPUFA*** | 23.62 (21.29-25.95) | 22.85 (19.79-25.91) | 26.95 (23.43-30.46) | * |

ΣPUFA: Sum of polyunsaturated fatty acid, ΣPUFA n-6 sum of omega 6 polyunsaturated fatty acid, ΣPUFA n-3: sum of omega 3 polyunsaturated fatty acid. ΣMUFA: sum of monounsaturated fatty acid, MUFA are reported in %.

The meat and poultry category (n=24) which includes beef, chicken, pork, turkey, and combinations of these meats, had the third-largest amount of total fat (values ranging from 0.60 to 33.13 g/100 g of product). The majority of UPFs showed a higher fat content compared to the values reported either in their nutritional label or website. Higher fat contents were observed in several dairy products containing butter as part of their use in their preparation process. Butter is the only processed food included in this study due to its extensive use in the manufacturing of several UPFs.

### 3.2 Saturated fatty acids (SFA)

Saturated fatty acid was the overall second major content of fatty acids between both FF and RTE meals with 35.57%, just behind the monounsaturated fatty acids (37.82%). The total SFA content was significantly different between RTE and FF with 40.88 and 29.88 g/100g of product, respectively. C16:00 was the predominant SFA in both RTE and FF meals with a mean of 21.74 g/100 g, followed by C18:00 with 7.61 g/100g. Regarding the length of the saturated chain in the fatty acid, long-chain fatty acid (LC-SFA) accounted for more than 90% of the total SFA Content in comparison with 5.3% of the medium-chain saturated fatty acid (MC-SFA). Dietary triacylglycerols are derived principally from two sources: animal fats and vegetable oils. Animal fats contain a high proportion of SFA as for butter, which SFA content mostly consists of 4 SFAs (C12:00, C14:00, C16:00, and C18:00) (Bobe et al., 2003; Ledoux et al., 2005; Lopes et al., 2019). On the other hand, vegetable oils have a higher proportion of unsaturated fatty acids. Since LCFA are hydrophobic nutrients, their intestinal absorption is a complex process. Fatty-acid chain length and unsaturation number influence fat absorption. In the human diet, approximately 95% of dietary lipids are triacylglycerols (TAG), mainly composed of long-chain fatty acids (LCFA, number of carbons >16). Medium-chain fatty acids (MCFA) are better absorbed than LCFA because they can be solubilized in the aqueous phase of the intestinal contents, absorbed bound to albumin, and transported to the liver by the portal vein (Wang et al., 2013). Further, the absorption in the stomach occurs after the hydrolysis of medium-chain triglycerides (MCT) by gastric lipase, enhancing their solubilization in the intestine, where they are absorbed bound to albumin and transported to the liver via the portal vein (Niot et al., 2009). LCFA exerts basic functions in the cell as membrane components, metabolic fuel, precursors of lipid mediators, regulators of ion channels, and modulators of gene expression (Linder & Deschenes, 2007). They participate in several post-translational protein modifications (e.g., palmitoylation) affecting their cellular functions. Moreover, dietary lipid fecal loss remains below 5% (w/w), still with a high fat intake in healthy individuals (Ramirez, Amate, & Gil, 2001). Hence, lipid uptake is not a rate-limiting step for intestinal fat absorption, and more attention should be paid to the meal size portion and fat source to maintain a healthy diet.

Interestingly, albeit not surprising considering the lipid needs for infants, infant formula has the overall highest SFA content from all UPFs with 56%. SFA content in infant formulas containing dairy as their major source of fat is significantly higher in comparison with those having plant oils as their main source (Kilvington, Barnaba, Rajasekaran, Laurens, & Gabriela Medina-Meza, 2021; Sun, Wei, Su, Zou, & Wang, 2018).

Even though the majority of SFA in both RTE and FF meals were straight chain even-numbered homologs (i.e., C12:00, C14:00, C16:00, etc.), C15:00, C17:00, and C23:00 were also detected in UPFs, being C15:00 the only statistically significant. Dairy foods, comprised of RTE items, contain the highest amount of C15:00 with 1.02 g/100g of product followed by the baby foods category. All type of cheeses (American, Cheddar, and Swiss) showed a high SFA content which agrees with the literature (Manuelian, Curro, Penasa, Cassandro, & De Marchi, 2017); except for the cream cheese, which literature values were not available to confirm their high SFA content. Mac & Cheese (boiled, prepared) showed a higher fat content than the Mac & Cheese prepared by microwave heating. These results demonstrate that cooking techniques and preparation are critical parameters that should be considered when evaluating the nutritional quality of foods. Total fat content for butter and ice cream agreed with previous studies (Nielsen, Olsen, Jensen, & Skibsted, 1996; Tavella et al., 2000) and the USDA food database (USDA, 2018).

The UPFs classified as “Others” (French fries, biscuits, hotcakes, mashed potato) contained 30.7% SFA, which was confirmed with their reference values in the USDA database (USDA, 2019r, USDA, 2019g, Huang et al., 2019). Palmitic acid (C16:0), lauric acid (C12:0), stearic acid (C18:0), and myristic acid (C14:0) were the four leading FA between the “Other” category.

RTE and FF meals are the UPFs with a major contribution of fat accumulation in the human body in the Western diet (Mohiuddin & Nasirullah, 2020), which is directly associated with different chronic diseases such as obesity, diabetes, hypertension, atherosclerosis, among others (Shori et al., 2017). Therefore, the quantification of total fat in RTE and FF meals is important to evaluate the actual dietary fat intake of adults and children who consume a large amount of UPFs.

### 3.2 Monounsaturated fatty acids (MUFA)

MUFA was the group with the highest content among all UPFs. FF values ranged from 40 to 45 (g/100 product) (**Table 3**) while RTE values range from 34 to 38 (g/100 product). It is worth mentioning that RTE meals contain higher percentages of SFA (40.88 g/100 of product) than the overall percentage of MUFA among UPFs. Conversely, the FF group contains the highest percentage of MUFA than the overall amount of both SFA and MUFA in all UPFs. Furthermore, the oleic acid (C18:1, cis-9, OA) was the dominant fatty acid in all UPFs with more than 90% of the total MUFA. OA not only provides energy but also reduces the melting point of triacylglycerides (Ramirez et al., 2001). Baby foods and meat & poultry were the categories with the higher content of OA, followed by seafood. The high level of OA on these meals may be due to the use of soybean and high oleic sunflower oil in their recipes. Palmitoleic acid (C16:1), oleic acid (C18:1, cis-9), and elaidic acid (C18:1, trans-9) were the second, third, and fourth most abundant MUFA in the meat & poultry category, respectively. Values are aligned with those reported previously (Haak, De Smet, Fremaut, Van Walleghem, & Raes, 2008; Karakok, Ozogul, Saler, & Ozogul, 2010) and with the information reported in their nutritional labels. Even though the majority of the MUFA reported in the USDA database were detected in our samples, some of them were present in small quantities in the USDA database and not detected in our samples and vice versa.

Twelve UPFs containing beef as their major component showed the highest fat content among the meat & poultry categories (**Table 4**). This abundant presence of fat in beef is also observed in other food items such as roasted rib (style: large end) which contains 26 g per serving size (3 oz) (USDA, 2019a), and also in ground beef which can contain a maximum 30% fat allowed by the USDA (USDA, 2019b). Previous studies in beef have reported SFA and MUFA as their dominating FAME group (Karakok et al., 2010; Wood, 1996). Beef with vegetables from a FF restaurant reported a saturated fat content of 21.4% on their website, which is slightly lower than our 26.94% of SFA, this confirms that our results could be more accurate.

**Table 4.**
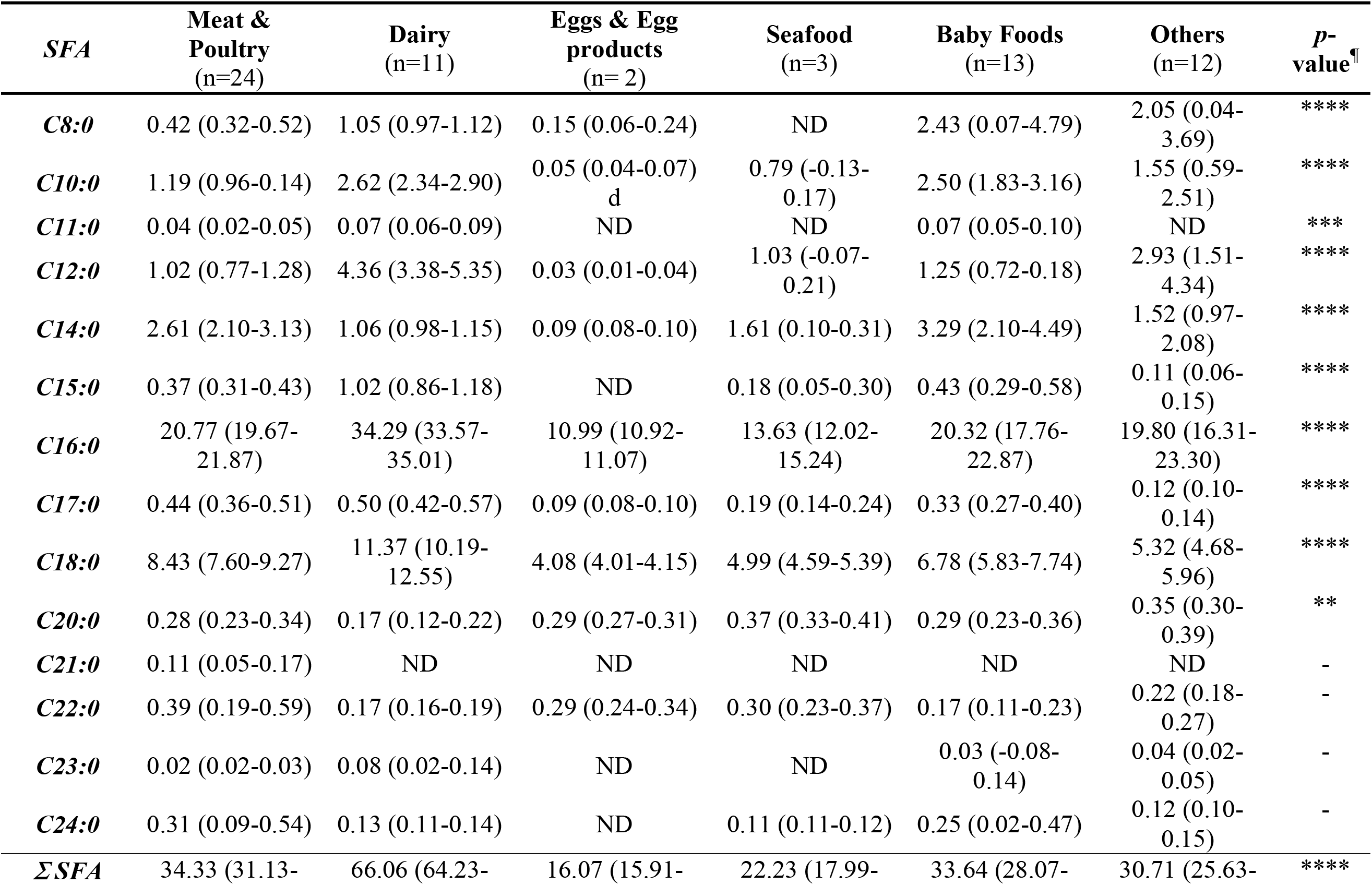
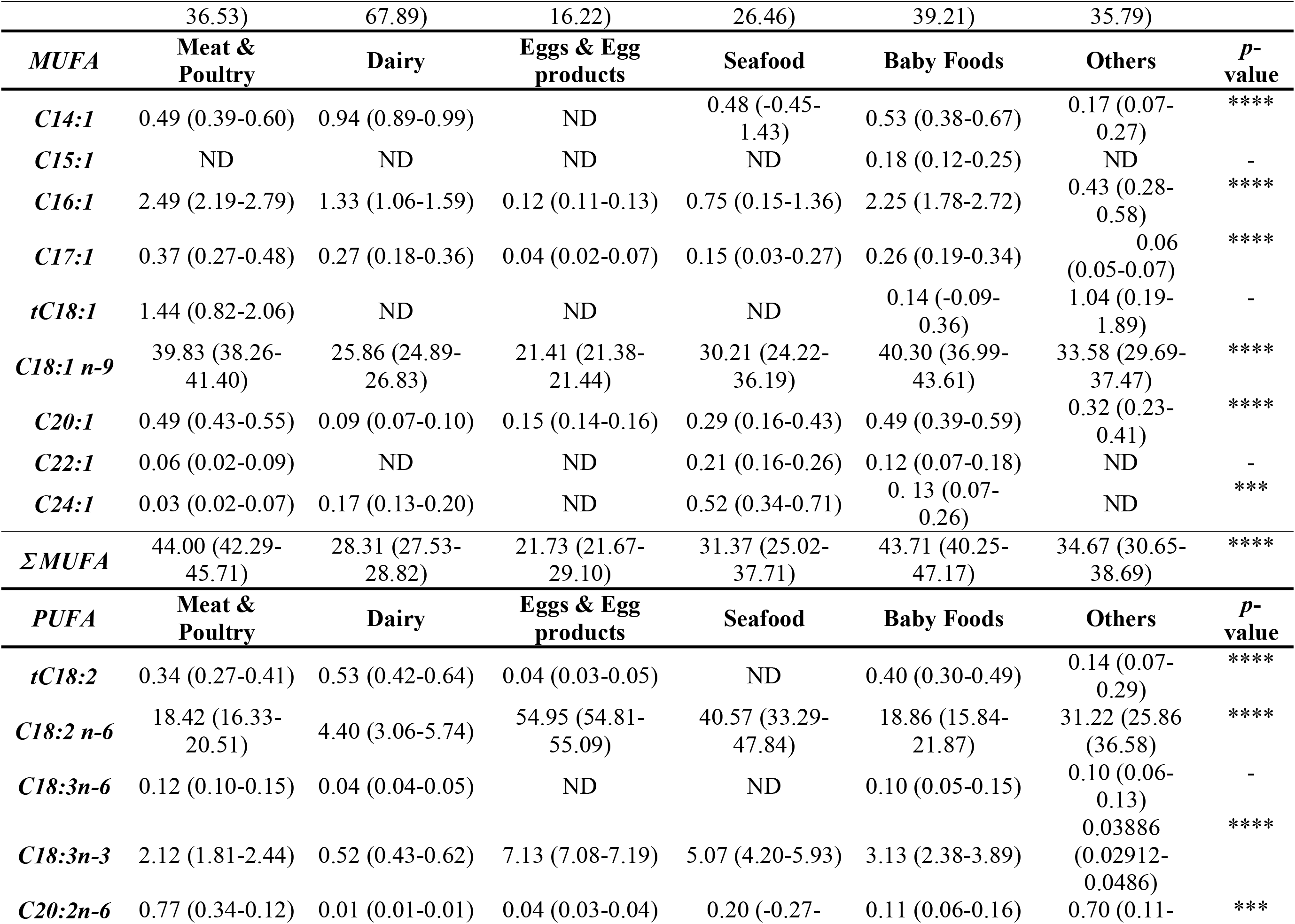
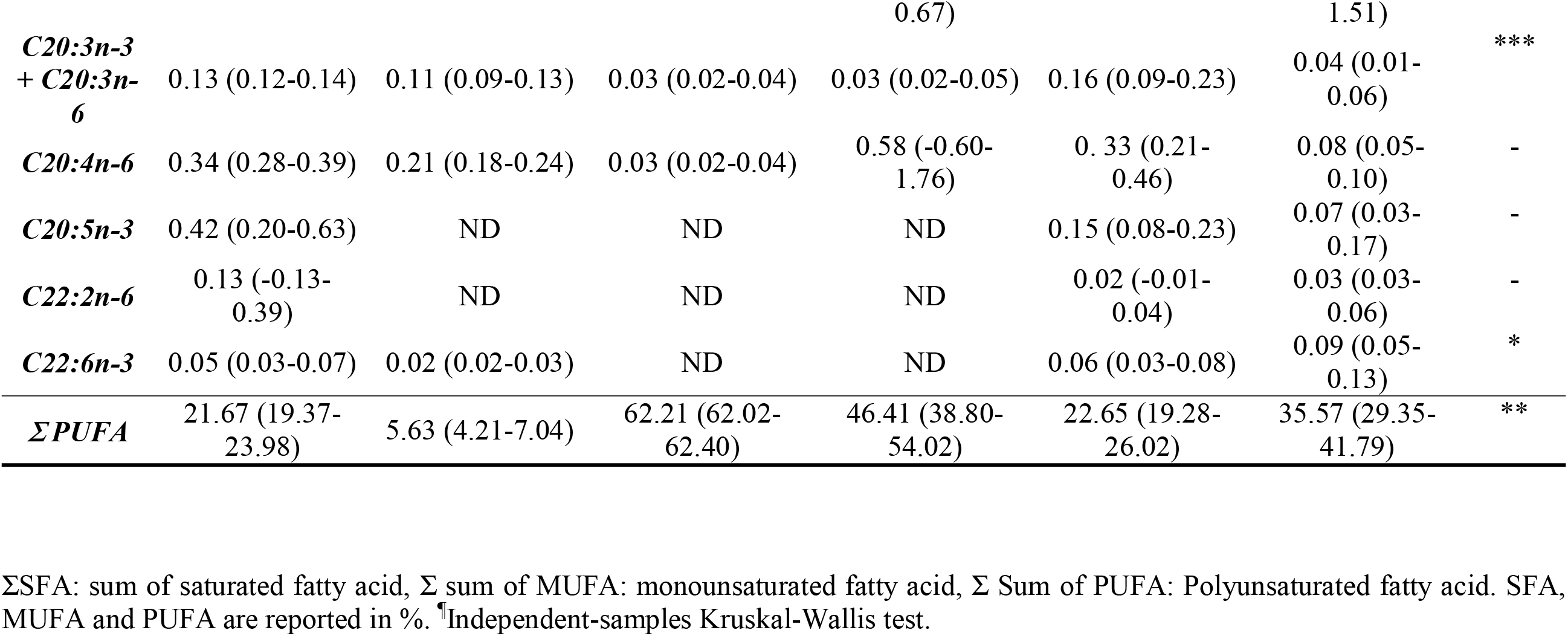
Fatty acid profile of UPFs by food category (mean and range).

Baby foods are RTE meals with the highest fat and MUFA contents, which are required to fulfill the infants’ growing needs. The main fat sources for baby foods are meat and poultry, chicken broth, and canola oil.

MUFA was dominated by oleic acid (C18:1, cis-9), palmitoleic acid (C16:1), and 11-eicosenoic acid (C20:1). PUFA in baby foods was led by linolenic acid, LA (C18:2cis), and γ-linolenic acid (GLA, C18:3n6). Lastly, less than 1% of the total fat contained trans fatty acids. Results agree with federal regulations about the total content of trans fatty acid in foodstuff (Albuquerque, Santos, Silva, Oliveira, & Costa, 2018).

### 3.3 Polyunsaturated fatty acids (PUFA)

Meals rich on PUFA are directly related to the use of vegetable oils and other ingredients such a fish, avocado, among others, which contain high PUFA levels. PUFA was the third most abundant group in all UPFs with no significant differences between FF and RTE. Linoleic acid (C18:2 n-6, LA) was the most abundant followed by α-linolenic acid (C18:3 n-3, LNA). The egg & egg’s derivatives category has the highest content of LA followed by seafood. These results agree with the literature (Bemrah, Sirot, Leblanc, & Volatier, 2009; Simopoulos & Salem, 1992). The main ingredient of macaroni salad is mayonnaise, and both have a similar fatty acid profile in agreement with those reported in literature (Tavella et al., 2000). Therefore, PUFA was also the predominant group in macaroni salad, and traces of trans fatty acid (18:2, trans-9,12) was detected just in mayonnaise.

Higher PUFA content was found in salad dressing, where results agree with previous studies that reported similar values (Jacobsen, 2015; Let, Jacobsen, & Meyer, 2007). It is worth mentioning that the effect of the cooking on FA profiles, for example, Mac & Cheese meals were cooked using 2 different cooking methods (boiled and microwaved, **Table S1)**. No reference values were found in both types of cooking methods employed in this study, however, the major ingredient on these meals is cheddar cheese which has been previously reported (Manuelian et al., 2017) and used for comparison purposes of this RTE. The microwaved mac & cheese showed a higher SFA content (55.6%) in comparison with the boiled sample (25.2 %), however the boiled sample has higher PUFAs content (52.5%) versus 17.52% of the microwave preparation, demonstrating the effect of temperature and type of cooking on variability on the nutritional content.

Seafood is well known to be a great source of LCFA, especially omega 3 long-chain PUFA (Abad, 2004; Bemrah et al., 2009; Costa et al., 2017; Neff, Bhavsar, Braekevelt, & Arts, 2014; Sirot, 2008). LC n−3 FA is primarily found in organisms that live in cold water. These organisms are especially dependent on the physicochemical properties of the FA. This is due to the high number of double bonds in the molecule lowering the melting point of the compounds, which means that, even at low temperatures, biological structures retain the fluidity necessary for life processes. Sources of LC n−3 FA with the greatest nutritional significance for humans are cold-water fish, such as salmon, mackerel, herring, and tuna. The USDA recommends fish consumption of at least 8 oz (227 g) seafood per week in individuals two years old and older (USDA, 2016). Seafood can be found in food retail (56%) and restaurants (31%); being shrimp, clams, salmon, cod, and Alaskan Pollock the most consumed seafood in the US (Love et al., 2020).

As expected, seafood meals contain the highest percentage of PUFA (46.41%) with minimal differences when comparing to previous literature values (Awogbemi, Onuh, & Inambao, 2019; Czech, Grela, & Ognik, 2015; Ekiz & Oz, 2019; Katan et al., 2020; Modzelewska-Kapituła, Pietrzak-Fiećko, Tkacz, Draszanowska, & Więk, 2019; Rant et al., 2019; Shan et al., 2019). For example, in fried shrimp from a FF restaurant, no C22:6 was detected as opposed to what its reference values state; and instead of C18:3n3, we were able to identify just C18:3n6 (Czech et al., 2015). Moreover, for the fish sandwich from another FF restaurant, PUFA was not the dominating FA group as stated in references because of the significant abundance of C18:2cis in our sample compared to the higher presence of C18:1cis reported by the USDA (USDA, 2020a). Results from RTE clam chowder soup are aligned with its reference values, however, a difference of 55.91% between PUFA and MUFA is reported in the USDA while our sample shows only a 1.05% difference. This suggests that different formulations for the same product may be available. RTE soups like the clam chowder is a canned product that usually needs reconstitution and warming steps before consumption. Differences during homemade preparation such as temperature changes, delay in warming, use of microwave vs boiling, etc. may increase the lipid oxidation process promoting autooxidation of lipid molecules which in turn, alters the overall fatty acid profile of the food item.

Differences in the PUFA content were observed between biscuits from two FF franchises. For example, EPA was present in O8-FF instead of ALA which was seen in O6-FF (**Supplementary Material – Table S1)**. This could be the result of the use of milk as part of the ingredients in O8-FF recipe which contains small amounts of EPA (USDA, 2018), in addition, to crosscontamination of this FA by the use of the same equipment for the cooking procedure of different food meals which could contain EPA. Lastly, even though 4 trans FAs were reported by USDA, no trans FA was detected in O8-FF. Overall, FA profile was similar to the one reported in the USDA database, and previous studies (Amrutha Kala, 2014; KFC, 2020; McDolnald’s, 2017; Rutkowska, Adamska, Sinkiewicz, & Białek, 2012; Smith, 1985), even though traces of trans FAs were reported by the USDA for French fries, none was detected in our sample. It is noteworthy to mention that the variability in frying oils used as part of the French fries cooking procedure in these 2 FF restaurants, may result in the presence of elaidic acid in item O9-FF, and its absence in O5-FF. As Smith and co-workers explained in their study (Song et al., 2015) the use of hydrogenated soybean oil for frying purposes caused the presence of high amounts of elaidic acid (C18:1, trans-9), while the use of an animal-vegetable mix shortening (corn oil and beef flavor) in O5-FF inhibited the formation of elaidic acid (C18:1, trans-9).

### 3.4 Correlations between macronutrients and price per serving on UPFs

One of the major objectives of this study was to evaluate correlations among evaluated parameters. Thus, we performed Spearman’s correlation among major FAME groups (SFA, MUFA, PUFA), macronutrients (sugars, fat), sodium, calories per serving (**Figure 2**), and price (total and per serving). Values of sugar and sodium were taken from the nutritional label, whereas fat content values were from our quantified fat determinations (**Table 1**).

**Figure 2.**
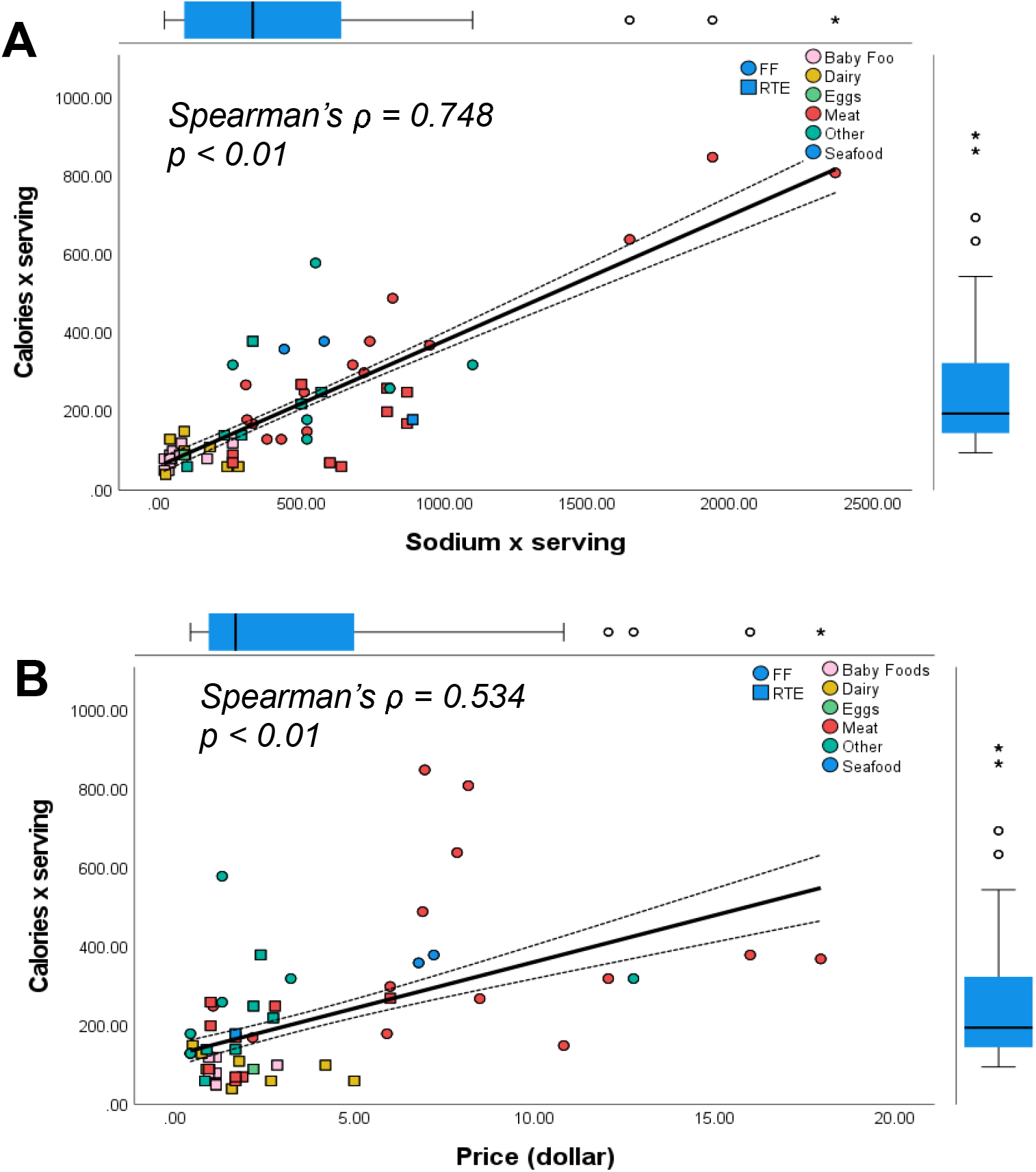
Correlations for price, calories per serving and sodium between RTE and FF and each food category.

When the overall UPFs parameters were considered, positive correlations (*ρ* = 0.748, *p* < 0.01) between calories per serving and sodium content were observed. The US Food and Drug Administration (FDA) estimates that Americans eat on average about 3,400 mg of sodium per day, while the recommendation from the Dietary Guidelines for Americans limits the sodium intake to less than 2,300 mg per day—that’s equal to about 1 teaspoon of salt (FDA, 2020b). Salt is widely present in the Western diet with over 70% of dietary sodium coming from UPFs. Particular attention must be given to the consumption of foods rich in salt which has been associated with the increased risk of developing hypertension, a major contributor of cardiometabolic disorders, kidney failure, and premature mortality (FDA, 2020a). As expected, inverse correlation were found between SFA and PUFA (*ρ* = -0.848, *p< 0*.*01*).

The most popular UPFs were used in this study, with prices ranging from 1 to 18 dollars per individual serving. The total price and calories per serving were also positively correlated (*ρ*= 0.534, *p* < 0.01). The price of the majority of the UPFs is below 5 dollars, having 2.5 dollars per serving as mode value. Calorie content ranges from few calories up to 800 in UPFs that contain meat as the major component. It is well known that UPFs are popular for being fast, cheap, and convenient meals. It is important to note that price and accessibility are critical factors for people facing food insecurity; which clearly may modify their dietary preferences and nutritional patterns. More attention should be paid to the relationship between price and serving size since cheaper meals are used to be also the most popular because of their big serving amount which attracts consumers looking for the most affordable and available meals.

## Conclusions

This study provides a systematic and novel assessment of the total fat, and fatty acid composition of the most popular UPFs in the US Mid-West area. Convenience and palatability are the main reasons to consume UPFs. However, nutritional quality and dietary patterns are jeopardized by prices and popularity of UPF meals resulting in a public health issue that should be addressed. The presented results revealed a positive trend in the use of plant-based oils in the elaboration of UPFs showing an increase of PUFA content when are cateogorized as RTE and FF; however, when UPFs are classified according to the fat source, dairy and baby foods are the group with less PUFA content. UPFs are greatly appreciated by children and teenagers, but also consumed by infants, which as a vulnerable population add additional concern from a public health point of view. The significance of this study is to provide new data about fat, salt, and sodium in the most consumed UPFs, with the opportunity to define priority interventions for a more advanced precision nutrition, especially for vulnerable populations.

Concerning the sugar, salt, and saturated fat content, it was noticed that new challenges have arisen, especially focusing on the food industry and policymakers. Some key changes may have a great impact such as reformulation of menu items, including more vegetable and fruit options as side and/o desserts, with the reduction of serving size especially in beverages, fatty sides, and desserts.

The gradual reduction of the aforementioned components in most of the food categories studied here should become a priority. It is possible to conclude that this subject remains present and new challenges are being pointed out by several national and international organizations aiming to improve the nutritional quality of ultra-processed foods.

## Data Availability

The data that support the findings of this study are available from the corresponding author, I.G.M.M., upon reasonable request.

## Acknowledgments

This study was funding by Center for Research Ingrediente Safety (CRIS) trough the GR100229 grant. USDA National Institute of Food and Agriculture, Hatch project MICL02526 to I.G.M.M. Authors are greateful with Nama Naseem, Nicole Urrea, and Lisa Zou for their assistance during lipid extraction.

