## Supplemental Information for "Evaluation of the Nutritional Quality of Ultra-Processed Foods (Ready to Eat + Fast Food): Fatty Acid Composition"

**Table S1.** Processed and Ultra-processed meals ID used in this study.

| ***Ready to Eat (RTE)*** | Dairy | **Sample ID** | ***Ultra-processed Foods (UPF)*** |
| --- | --- | --- | --- |
|  |  | D1 - RTE | American Cheese – Happy Farms |
|  |  | D2 - RTE | Cheddar cheese – Happy Farms |
|  |  | D3 - RTE | Margarine (regular, not low-fat, salted) – Countryside Creamery |
|  |  | *D4 – RTE | Butter – Praire Farms |
|  |  | D5 - RTE | Cream (half & half) -Meijer |
|  |  | D6 - RTE | Swiss cheese - Kroger |
|  |  | D7 - RTE | Cream cheese – Happy Farms |
|  |  | D8 - RTE | Ice cream (regular, not low-fat, vanilla) - Purple Cow |
|  |  | D9 - RTE | Yogurt (low-fat, fruit flavored) - Yoplait |
|  |  | D10 - RTE | Chocolate milk - Nesquick |
|  |  | D11-RTE | Infant formula |
|  | Meat & Poultry | MP1 - RTE | Bologna - Eckrich |
|  |  | MP2 - RTE | Salami – Oscar Mayer |
|  |  | MP3 - RTE | Soup bean w/bacon/pork (canned, prepared w/water) - Campbell's |
|  |  | MP4 – RTE | Chili con carne w/beans (canned) - Campbell's |
|  |  | MP5 – RTE | Lasagna w/meat (frozen, heated) – Michael Angelo’s |
|  |  | MP6 – RTE | Chicken noodle soup - Kroger |
|  |  | MP7 – RTE | Beef and vegetables soup – Kroger |
|  |  | MP8 – RTE | Mini Ravioli - Chef Boyardee |
|  |  | MP9 - RTE | Spaghetti - Chef Boyardee |
|  | Seafood | S1 - RTE | Clam chowder (New England, canned, prep w/ whole milk) – Kroger |
|  | Eggs & egg derivatives | E1 - RTE | Mayonnaise (regular, bottled) – Hellmann’s |
|  |  | E2 - RTE | Macaroni salad (from grocery/deli) - Meijer |
|  | Baby food | BF1 - RTE | Baby food - beef and broth/gravy – Beech Nut |
|  |  | BF2 - RTE | Baby food – chicken and broth/gravy - Gerber |
|  |  | BF3 - RTE | Baby food - vegetables and beef - Gerber |
|  |  | BF4 - RTE | Baby food - vegetables and chicken - Gerber |
|  |  | BF5 - RTE | Baby food - chicken noodle dinner - Gerber |
|  |  | BF6 - RTE | Baby food - macaroni, tomato and cheese - Gerber |
|  |  | BF7 - RTE | Baby food - turkey and rice - Gerber |
|  |  | BF8 - RTE | Baby food – turkey and broth/gravy – Beech Nut |
|  |  | BF9 - RTE | Baby food – fruit yogurt - Gerber |
|  |  | BF10 - RTE | Baby food – chicken with rice - Gerber |
|  |  | BF11 - RTE | Baby food – vegetables and turkey - Gerber |
|  |  | BF12 - RTE | Baby food – macaroni and cheese with vegetables - Gerber |
|  |  | BF13 - RTE | Pasta pick-ups (cheese ravioli) - Gerber |
|  | Others | O1 - RTE | Popcorn w/butter (microwave) - Kroger |
|  |  | O2 - RTE | Salad dressing (creamy/buttermilk type, regular) – Aldi’s Tuscan Garden |
|  |  | O3 - RTE | Macaroni & cheese (boiled) - Kraft |
|  |  | O4 - RTE | Macaroni & cheese (microwaved) - Kraft |
| ***Fast Food (FF*** | Meat & Poultry | MP10 - FF | Hamburger on bun – McDonald’s |
|  |  | MP11 - FF | Chicken nuggets – McDonald’s |
|  |  | MP12 - FF | Cheeseburger on bun – McDonald’s |
|  |  | MP13 - FF | Steak tacos w/beans, lettuce, rice and cheese - Chipotle |
|  |  | MP14 - FF | Cheese and chicken quesadilla - Chipotle |
|  |  | MP15 - FF | Chicken burrito w/lettuce, cheese, pico - Chipotle |
|  |  | MP16 - FF | Chicken drumstick - KCF |
|  |  | MP17 - FF | Chicken wing - KFC |
|  |  | MP18 - FF | Beef w/vegetables - Panda Express |
|  |  | MP19 - FF | Chicken w/vegetables - Panda Express |
|  |  | MP20 - FF | Chicken filet - (broiled sandwich) – Chick Fil’A |
|  |  | MP21 - FF | Roast beef, ham & Provolone - Jimmy Johns |
|  |  | MP22 - FF | Sliced turkey and bacon - Jimmy Johns |
|  |  | MP23 - FF | Supreme pizza - Marco's Pizza |
|  |  | MP24 - FF | Pepperoni pizza, hand tossed - Domino's |
|  | Seafood | S2 - FF | Fish sandwich on bun – McDonald’s |
|  |  | S3 - FF | Fried Shrimp – Panda Express |
|  | Others | O5 - FF | French-Fries – McDonald’s |
|  |  | O6 - FF | McDonald's Biscuit - Big Breakfast |
|  |  | O7 - FF | McDonald's Hotcakes - Big Breakfast |
|  |  | O8 - FF | Biscuit - KFC |
|  |  | O9 - FF | French Fries - KFC |
|  |  | O10 - FF | Mashed potato - KFC |

*The only processed food included in this study.
